# SARS-CoV-2 convalescence and hybrid immunity elicits mucosal immune responses

**DOI:** 10.1101/2023.03.24.23287677

**Authors:** Olha Puhach, Mathilde Bellon, Kenneth Adea, Meriem Bekliz, Krisztina Hosszu-Fellous, Pascale Sattonnet, Sophie Coudurier-Boeuf, Isabelle Arm-Vernez, Laurent Kaiser, Isabella Eckerle, Benjamin Meyer

## Abstract

Mucosal antibodies play a key role in the protection against SARS-CoV-2 infection in the upper respiratory tract, and potentially in limiting virus replication and therefore onward transmission. While systemic immunity to SARS-CoV-2 is well understood, little is known about the antibodies present on the nasal mucosal surfaces.

In this study, we evaluated SARS-CoV-2 mucosal antibodies in response to infection, vaccination, or a combination of both. Paired nasal fluid and serum samples were collected from 136 individuals, which include convalescent, vaccinated, or breakthrough infections.

We detected a high correlation between IgG responses in serum and nasal fluids, which were higher in both compartments in vaccinated compared to convalescent participants. Contrary, nasal and systemic SARS-CoV-2 IgA responses were weakly correlated, indicating a compartmentalization between the local and systemic IgA responses. SARS-CoV-2 secretory component IgA (s-IgA) antibodies, present exclusively on mucosal surfaces, were detected in the nasal fluid only in a minority of vaccinated subjects and were significantly higher in previously infected individuals. s-IgA binding antibodies showed significant correlation with neutralizing activity of nasal fluids against SARS-CoV-2 ancestral B.1 and Omicron-BA.5 variant, indicating that s-IgA is the crucial contributor to neutralization in the nasal mucosa. Neutralization against both SARS-CoV-2 strains was higher in the mucosa of subjects with previous SARS-CoV-2 infections compared to vaccinated participants.

In summary, we demonstrate that currently available vaccines elicit strong systemic antibody responses, but SARS-CoV-2 infection generates more potent binding and neutralizing mucosal antibodies. Our results support the importance to develop SARS-CoV-2 vaccines that elicit mucosal antibodies.

**One Sentence Summary:** SARS-CoV-2 infection or combination of infection and vaccination (hybrid immunity) elicit binding and functional mucosal antibody responses superior of those after systemic vaccination.

## Introduction

Less than one year after the emergence of severe acute respiratory syndrome coronavirus 2 (SARS-CoV-2), the causative agent of coronavirus disease 2019 (COVID-19), highly effective COVID-19 vaccines were licensed^1,2^. To date, more than 13 billion doses of COVID-19 vaccines have been administered worldwide, significantly reducing the number of severe diseases and hospitalizations^3^. However, breakthrough infections have been reported even for SARS-CoV-2 variants that are antigenically similar to the vaccine strain and more frequently since the emergence of the Omicron variant with its efficient immune evasion properties^4,5^. These results indicate that current vaccines can only temporarily and incompletely reduce the risk of upper respiratory tract (URT) infections^6,7^. In contrast, studies in mice and rhesus macaques demonstrated that intranasal immunization leads to complete protection against SARS-CoV-2 infection of the URT, resulting in reduced onward transmission^8,9^. In humans, the level of mucosal IgA antibodies, elicited mainly by previous SARS-CoV-2 infection, was associated with protection from infection with Omicron subvariants^10^. However, the origin of these mucosal IgA antibodies as well as their functional properties have not been investigated in detail.

The URT harbours a distinct part of the immune system called the mucosa associated lymphatic tissue (MALT), where local plasma cells secrete multimeric IgA (mainly dimers but also trimers and tetramers), which is transported across the mucosal epithelium by the polymeric immunoglobulin receptor (pIgR). During this process the extracellular part of the pIgR, called the secretory component (SC) is bound to the multimeric IgA forming secretory IgA (s-IgA). The main function of s-IgA antibodies is neutralization^11^. In particular, s-IgA dimers were shown to neutralize SARS-CoV-2 on average 15 times more potently than IgA monomers present in serum^12^. Mucosal IgG is either locally produced by B cells in the lamina propria and transported across the epithelium by neonatal Fc receptors, or excreted as a transudate from serum together with monomeric IgA antibodies^13^. While potent systemic IgA and IgG responses are shown to be induced by infection and vaccination, mucosal antibody responses are mainly induced by infection^14^. Notably, some studies demonstrated that virus-specific mucosal IgA antibodies were found in some seronegative COVID-19 patients, suggesting a discrepancy between local and systemic antibody immune responses^15,16^. In convalescent patients higher SARS-CoV-2-specific mucosal antibodies were associated with lower viral loads and more efficient symptom resolution^17^; these antibody responses remained detectable for at least 3 months post infection in saliva^18^, and up to 9 month post infection in nasal secretions^17,19^.

Vaccination was shown to significantly reduce infectious viral loads in breakthrough infections with the Alpha variant, but the effect was weaker for breakthrough infections with Delta ^20,21^, and even less potent for the Omicron BA.1 variant, where the decrease of viral loads was shown only after boosting with a 3^rd^ vaccine dose^22,23^. However, it remains unclear whether this reduction is caused by mucosal or systemic immunity. Even though s-IgA responses were detectable in saliva of some of the vaccinated subjects^24^, these responses were significantly lower in comparison to previously infected individuals^25,26^. Additionally, most virus-specific antibody titers in saliva and their neutralizing capacity significantly decayed six months post vaccination^25^. One study showed that mucosal antibody responses induced by vaccination were low or undetectable, but Delta breakthrough infections led to significant increases of antibody titres in saliva^27^. According to one study, previous infection, but not vaccination, induced strong IgA responses and detectable neutralizing titers against Omicron subvariants in the nasal mucosa^28^.

In this study we characterized mucosal antibody responses in the URT of convalescent, vaccinated and subjects with vaccine breakthrough infections. We quantified and compared the levels of mucosal and systemic IgA and IgG responses in these groups as well as analyzed the levels of neutralizing antibodies in the nasal lining fluid (NLF).

## Materials and methods

### Study design and participants

The study was approved by the Cantonal Ethics Committee at the University Hospital of Geneva (CCER no. 2020-02323). All study participants provided written informed consent. NLF and serum samples were collected during a single visit from all the participants from September 2021 to June 2022. The samples were collected from healthy volunteers with no symptoms of respiratory illness. All the sampling procedures were performed by trained healthcare professionals. The information about previous infections (date of infection confirmed by RT-PCR or rapid antigenic test) and vaccination (date of vaccination, number of vaccine doses and the vaccine manufacturer) was collected using a questionnaire during the visit.

NLF samples were collected using the Nasosorption™ FX·i nasal sampling device (Hunt Developments, UK) as previously described^29^. Briefly, a synthetic absorptive matrix (SAM) strip was inserted in the nostril of the participant against the inferior turbinate. After pressing on the side of the nostril for one minute, the SAM strip was removed and placed in the collection tube. The samples were stored at −80°C before processing. Nasosorption devices were thawed on ice, the SAM was removed and placed in the collection tube with 300 μl of elution buffer (PBS/1% BSA). Following incubation for 10 minutes at room temperature, the SAM was placed on the spin-X filter Eppendorf tube and centrifuged at 16,000 g for 10 min at 4 °C. Eluted liquid was aliquoted and kept at −80 °C until further use.

### Immunoassays

#### Roche Elecsys anti-SARS-CoV-2 S RBD and Roche Elecsys

Elecsys anti-SARS-CoV-2 S and anti-SARS-CoV-2 were used to determine the levels of SARS-CoV-2 specific RBD and nucleoprotein antibodies respectively. Antibody measurements were performed on the cobas e801 analyser (Roche Diagnostics, Rotkreuz, Switzerland) in the clinical laboratory of the University Hospital of Geneva. For anti-RBD antibodies, results are reported as concentrations (U/mL) and positivity was determined by using the manufacturer’s cut-off >0.8 U/mL. For anti-NP antibodies, the positivity was determined with a cut-off index (COI), where COI ≥1.0 is defined as positive.

#### Recombinant proteins

Full-length trimerized SARS-CoV-2 spike (triS) was provided by the EPFL protein production facility. Nucleoprotein (NP) was obtained from Prospec Bio.

#### Human total IgA ELISA

The levels of total IgA in NLF were measured by IgA Human Uncoated ELISA Kit (Catalog # 88-50600, Invitrogen) following the manufacturer’s instructions. NLF samples were diluted to 1:1000 and 1:10000 in assay buffer prior to measurement. The standard curve was generated from recombinant human IgA using four-parameter logistic (4PL) regression model. The relative IgA concentration (ng/mL) of test samples was determined according to the dynamic range of the standard curve by interpolating the concentration of the standards that correspond to the absorbance value. To normalize all SARS-CoV-2 IgA levels in NLF samples to total IgA, a correction factor calculated by dividing the mean of total IgA for all samples by the sample’s total IgA was applied to all the NLF samples.

#### IgA and IgG Anti-SARS-CoV-2 triS ELISA

Maxisorp plates (Thermo Fisher Scientific) were coated with SARS-CoV-2 full triS protein diluted in Phosphate-Buffered Saline (Thermo Fisher Scientific) at a concentration of 2µg/ml. 50 µl of diluted antigen was added to each well and incubated overnight at 4°C. To control for unspecific binding half of the plate was coated with PBS only. Following three washes with washing buffer (PBS/0.1% tween-20), plates were blocked for one hour at 37°C with assay buffer (PBS/1% BSA/0.1% tween-20). Human IgA SARS-CoV-2 S monoclonal antibodies (IgA1 AR222, Geneva Antibody Facility) were serially diluted in assay buffer (3-fold serial dilutions from 300 ng/ml to 0.41 ng/ml) and added to each plate to generate a relative IgA anti-S standard curve. Human IgG SARS-CoV-2 S monoclonal antibody (IgG1 AR222, Geneva Antibody Facility) (3-fold serial dilutions from 100ng/ml to 4.6 pg/ml) was used to generate IgG anti-S standard curve. For IgA anti-SARS-CoV-2 tris ELISA, 3-fold dilutions from 1:5 to 1:45 in assay buffer prepared for NLF and 1:100, 1:900, and 1:2700 dilutions in assay buffer prepared for serum samples). For IgG anti-SARS-CoV-2 triS ELISA, 9-fold dilutions were prepared for serum (1:100 to 1:24300) and 3-fold dilutions were prepared for NLF (1:15 to 1:45). 50 µl of standard and sample dilutions in duplicates were added to coated and uncoated wells and incubated for 1 hour at 37°C. Plates were washed four times and 50 µl of Peroxidase AffiniPure Goat Anti-Human Serum IgA (Cat. # 109-035-011, Jackson ImmunoResearch) for IgA ELISA or Peroxidase AffiniPure F(ab’)₂ Fragment Goat Anti-Human IgG (109-036-098, Jackson) antibodies for IgG ELISA at a 1:5000 dilution were added to all wells. After 1 hour of incubation at 37°C and four washes and plates were developed with 3,3’,5,5’Tetramethylbenzidine (TMB) (Sigma-Aldrich) for 20 minutes in the dark. The reactions were stopped with 1N sulfuric acid. The developed plates were read at 450 nm wavelength. The absorbance values measured from uncoated wells were subtracted from values obtained from antigen-coated wells. The SARS-CoV-2 IgA standard curve was generated from the human IgA SARS-CoV-2 S mAb using a 4PL regression model. The relative anti-S IgA concentrations (ng/mL) of test samples were determined by interpolation of optical density values on a standard curve. The mean + 3SD of eleven negative samples was used to set a cut-off for anti-SARS-CoV-2 IgA and IgG in NLF. The mean + 3SD of 56 or 48 pre-pandemic samples was used to set a cut-off in serum for IgA anti-SARS-CoV-2 or IgG anti-SARS-CoV-2 respectively.

#### Multiplex immunoassay

For s-IgA analysis in NLF, a fluorescent-bead-based multiplex immunoassay was developed. Full-length His-tagged SARS-CoV-2 Spike and Nucleoprotein were each coupled to MagPlex carboxylated polystyrene microparticles using xMAP ® Antibody Coupling Kit (Luminex) following manufacturer’s instructions. Antigen-conjugated microspheres were resuspended at a concentration of 1×10^4^ /ml and incubated in assay buffer (PBS/1% BSA/0.1% tween-20) for 1 hour at room temperature. A total of 4 NLF samples collected from a subject with previously confirmed SARS-CoV-2 infection were pooled together and used to create standard curves for SARS-CoV-2 triS and NP s-IgA. 3-fold serial dilutions were performed for NLF samples (1:3 to 1:81 in assay buffer). Diluted samples and standards were incubated with antigen-coupled microspheres for 2 hours while shaking at 800 rpm. Following 3 washes with PBS/0.1% BSA/0.1% tween-20, mouse anti-Human IgA secretory component antibodies (Catalog # ABIN6155159, Antibodies online) was added to microspheres and incubated for 1 hour at room temperature. Following three washes the data was acquired on Bio-Plex® MAGPIX™ Multiplex Reader. MFI was converted to arbitrary units (AU/ml) by interpolation from a log-4PL-parameter logistic standard curve (3000 AU/ml to 4.12 AU/ml). Samples with values of less than 12 AU/ml were considered negative and an arbitrary value of 6 AU/ml was assigned to these samples. The mean + 3SD of eleven negative samples was used to set a cut-off for NLF.

#### Viruses and cells

Vero E6-TMPRSS cells (Catalog # 100978, National Institute for Biological Standards and Controls) were cultured in complete DMEM GlutaMAX medium supplemented with 10% FBS, 1× non-essential amino acids and 1% antibiotics (penicillin–streptomycin) (all reagents from Gibco).

All SARS-CoV-2 viruses used in this study were isolated from residual nasopharyngeal swabs collected from patients at the University Hospital of Geneva under general informed consent that allows the usage of anonymized left-over materials. All patient specimens from which isolates were obtained were fully sequenced. The SARS-CoV-2 B.1 variant was isolated and propagated on Vero-E6 cells. The Omicron-BA.5 variant was primarily isolated on Vero-TMPRSS cells, then transferred to Vero-E6 for generation of virus stock. All virus stocks were titrated on Vero E6-TMPRSS cells and fully sequenced. All infection experiments were performed under Biosafety Level 3 conditions.

#### Focus Reduction Neutralization Assays

Serially diluted NLF and SARS-CoV-2 (50 focus-forming units) were combined in serum-free Opti-Pro medium (Gibco) and DMEM + 1%FBS (Corning Cellgro) and incubated at 37°C for one hour. The antibody-virus mixture was added to a monolayer of Vero E6-TMPRSS cells and incubated at 37°C for one hour. After 1 hour at 37 °C, the media were removed, and pre-warmed medium mixed with 2.4% Avicel (DuPont) at a 1:1 ratio was overlaid. Plates were incubated at 37 °C for 24 hours and then fixed using 6% paraformaldehyde for 1 hour at room temperature. The staining for SARS-CoV-2 nucleocapsid protein was done as described previously^22^.

The 50% reduction endpoint titers (FRNT_50_) were calculated by fitting a 4-PL logistics curve with variable slope to the number of foci of each NLF using GraphPad Prism version 9.1.0. If the extrapolation reached a titer below 0.5, the value of 0.5 was attributed to the sample.

### Statistical analysis

Data collection was done using Excel 2019. All statistical analyses were performed using Prism version 9.3.1 (GraphPad). All IgA and IgG antibody titers were log_10_ transformed, and samples with no detectable antibodies were set to 1 ng/ml for the purpose of analysis. Differences between antibody titers between the different groups were analyzed with one-way ANOVA with Tukey test. Correlations between antibody titers were analyzed using Pearson’s rank test.

## Data Availability

All data produced in the present study are available upon reasonable request to the authors

## Abbreviations

NLF: nasal lining fluid
triS: trimeric Spike
NP: Nucleoprotein

## Results

### Study design and participants

In this study we determined the quantity and quality of SARS-CoV-2-specific mucosal antibodies in individuals that have only been vaccinated or infected, or have hybrid immunity and compared them to the systemic responses. A total of 143 adults were recruited between September 2021 and June 2022. Paired nasal lining fluid (NLF) and serum samples were collected during a single visit from all study participants. 11 participants, which had not been vaccinated and never tested positive for SARS-CoV-2, served as a negative control group. 29 and 25 participants had been vaccinated with 2 or 3 doses, respectively, but never tested positive for SARS-CoV-2. 21 participants had been tested positive for SARS-CoV-2 previously, but were not vaccinated. 31 and 26 participants tested positive after having received 1/2 or 3 doses of COVID-19 vaccine, respectively. All groups had a similar median age ranging between 28.5 and 39 years. Percentage of female participants was slightly higher in the negative, convalescent and vaccinated (2 doses) groups. All participants reported mild to moderated disease at the time of infection. We used days since the last immune response (DLIR) either elicited by infection or vaccination to compare the different groups. Median DLIR was 68 days for convalescent, 176 and 70 days for individuals vaccinated with 2 or 3 doses, respectively, and 44 or 47.5 days for vaccine breakthrough after 1/2 or 3 doses respectively (**Table 1**). Sequence information to determine the infecting variant was only available for a minority of participants. Nevertheless, in case only one variant circulated at the time of the positive test, we assumed that this variant infected the participant. If more than one variant circulated at the time, we marked the infecting variant as unknown. More details on the type of vaccinations received and the time between sampling, vaccination and infection are shown in **Table S1**.

**Table 1:** Participants characteristics

| Table 1: Participants characteristics |  |  |  |  |  |  |
| --- | --- | --- | --- | --- | --- | --- |
| Group | Negative | Vaccinated (2 doses) | Vaccinated (3 doses) | Convalescent | Hybrid Immunity (1 or 2 doses) | Hybrid Immunity (3 doses) |
| Group description | Non-vaccinated, never tested positive, NP negative | Never tested positive, anti-NP negative, anti-S positive | Never tested positive | Non-vaccinated, tested positive | Infection after 1 or 2 doses of vaccine | Infection after 3 doses of vaccine |
| Number of patients | 11 | 29 | 25 | 21 | 31 | 26 |
| Age: Median (range) | 28.5 (24-46) | 33 (24-59) | 37.5 (24 -76) | 32 (23-64) | 39 (25-59) | 37.5 (23-55) |
| Sex: Females (%) | 8 (72.7 %) | 20 (69 %) | 13 (52 %) | 16 (76.2 %) | 16 (51.6%) | 15 (57.7 %) |
| No of participants with >1 infection | na | na | na | 2 | 8 | 3 |
| Days since last immune response, days, median (IQR) | na | 176 (163 – 210) | 70 (50 - 123) | 68 (38.25 - 364.75) | 44 (29.5 - 54) | 47.5 (38.25 - 56.25) |
| Nr. of subjects with unknown date of infection | na | na | na | 3 | - | - |

To ensure that participants were assigned to the correct group, we tested serum samples for the presence of SARS-CoV-2-specific anti-nucleoprotein (NP) and receptor-binding domain (RBD) antibodies using the Roche Elecsys N and S assays. Highest anti-RBD serum responses were detected in participants vaccinated with 3 doses and with hybrid immunity, irrespective of the number of vaccine doses received. Participants vaccinated with 2 doses or convalescent individuals had lower anti-RBD titers, while no anti-RBD antibodies were detected in the negative group (**Figure S1A**). Anti-NP responses were only detected in convalescent individuals or participants with hybrid immunity, but not in negative or vaccinated participants confirming the absence of previous infection in these groups (**Figure S1B**). Therefore, we used samples from the negative cohort to determine the assay background and define the cut-offs of positivity for antibody responses in NLF. Since IgA concentrations in NLF might vary between individuals and between sampling we measured the level of total IgA. We found no significant differences of total IgA in the NLF of all groups compared to the negative control group, confirming that sampling of NLF by SAM strips leads to low sampling variability (**Figure S1C**). Nevertheless, to avoid a possible sampling bias on individual level, we used the levels of total IgA to normalize the levels of SARS-CoV-2-specific IgA and s-IgA antibodies in NLF.

### Distinct nasal and systemic IgA responses

First, we investigated whether there are differences between antibodies present at the nasal mucosa and those circulating the blood. Therefore, we measured the levels of IgA and IgG antibodies responses to trimeric spike (triS) protein of SARS-CoV-2 (strain: Wuhan-HU-1) in serum and NLF using an ELISA. We detected only a low correlation between levels of anti-triS IgA antibodies in serum and NLF (Pearson r=0.23, p=0.0082), but a high correlation between anti-triS IgG responses in serum and NLF (Pearson r=0.75, p<0.0001) **(Figure 1A and B)**. These findings indicate that while SARS-CoV-2 IgG mucosal and systemic responses are highly comparable, there is a compartmentalization between IgA responses in mucosa and serum.

**Figure 1.**
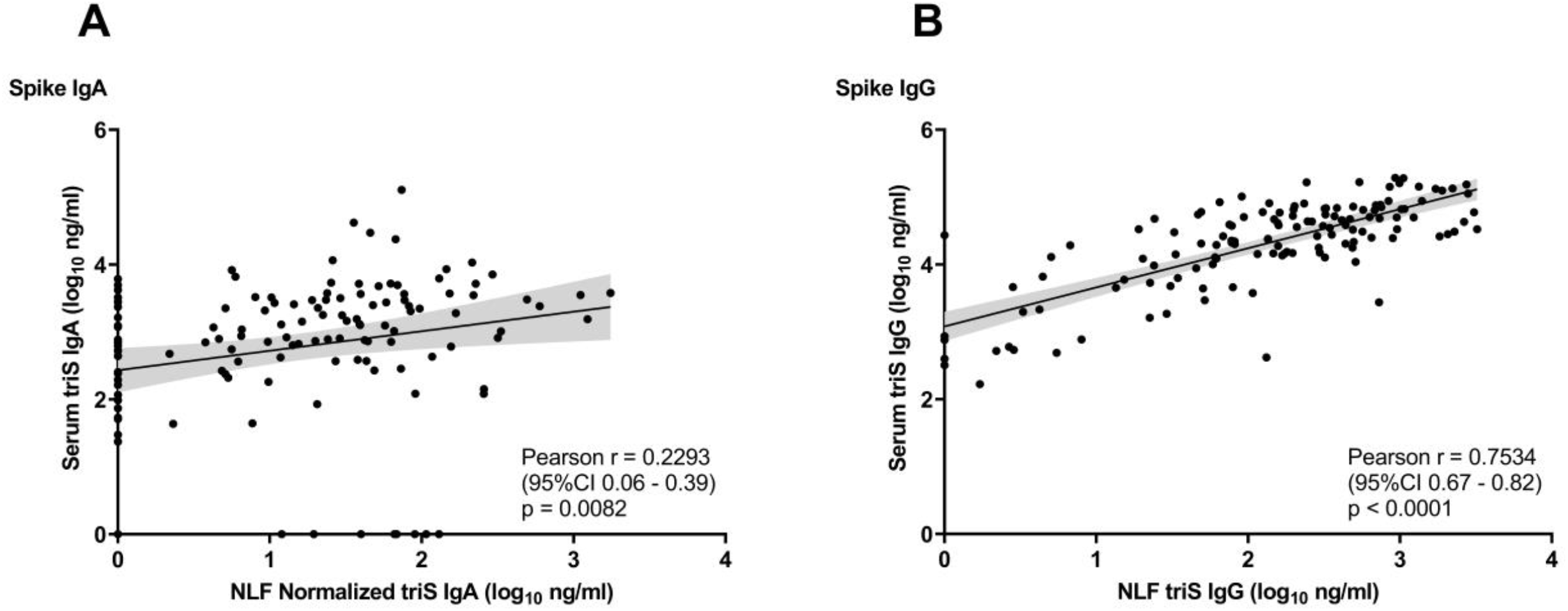
Correlation of SARS-CoV-2 IgA and IgG antibody responses in NLF and serum. A,B. Correlation between anti-triS serum and NLF IgA (A) and IgG (B) antibody titers (log_10_ ng/ml) of samples collected from convalescent, vaccinated and subjects with hybrid immunity. NLF triS IgA titers normalized to the levels of total IgA from the same sample. Pearson correlation coefficient and p values are shown.

### SARS-CoV-2 mucosal antibodies in response to infection and vaccination

Next, we investigated the level of mucosal antibodies induced by vaccination or infection by analyzing anti-triS IgA responses in NLF of vaccinated and convalescent subjects. In most of vaccinated subjects, NLF IgA responses were below the cut-off of positivity (21 out of 29 double-vaccinated and 15 of 25 in triple-vaccinated below the cut-off) and there was no difference if participants had received 2 or 3 doses despite the higher median DLIR in double-vaccinated individuals (176 vs 70 days). In contrast, positive IgA responses were detected in NLF of most of the convalescent individuals (16 out of 21 above cut-off), and they were significantly higher in comparison to vaccinated subjects (**Figure 2A**). Conversely, anti-triS IgA responses were moderately higher in serum of vaccinated subjects that had received 2 or 3 doses compared to the convalescent group, but the difference was not significant (**Figure 2B**).

**Figure 2.**
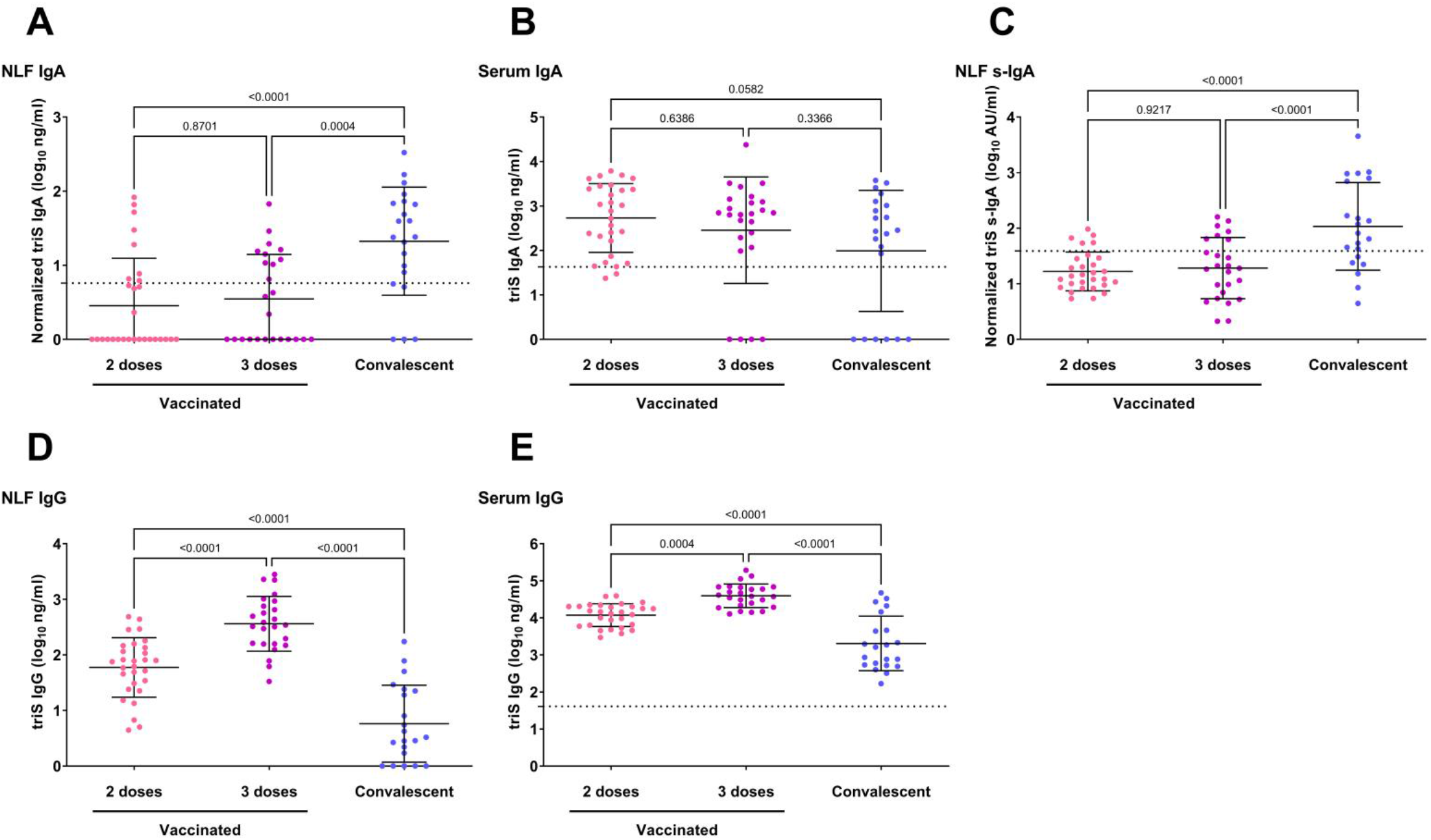
Mucosal and serum antibody responses in subjects with previous infection compared to vaccinated only. A, B. Anti-triS IgA (log_10_ ng/ml) measured in NLF (A) and serum (B) of vaccinated with 2 or 3 doses, or convalescent. (C) Anti-triS s-IgA (log_10_ AU/ml) antibody titers in NLF of vaccinated with 2 or 3 doses, or convalescent. D,E. Anti-triS IgG (log_10_ ng/ml) measured in NLF (D) and serum (E) of vaccinated with 2 or 3 doses, or convalescent. One-way ANOVA with Tukey test was used to determine differences of means, p values are shown above brackets. The mean + 3SD of 11 negative samples was used to set a cut-off for anti-triS IgA, s-IgA and IgG in NLF. The mean + 3SD of 56 pre-pandemic serum samples was used to set a cut-off for anti-triS IgA in serum samples. The mean + 3SD of 48 pre-pandemic serum samples was used to set a cut-off for anti-triS IgG in serum samples.

To exclude that the observed differences in IgA are biased by exudate of serum IgA, we analyzed the levels of locally-produced anti-triS s-IgA in NLF. Notably, we observed a strong and significant correlation between anti-triS IgA and anti-triS s-IgA responses in NLF (Pearson r=0.64, p<0.0001) (**Supplementary figure 2**) indicating that the majority of IgA measured in NLF is locally produced s-IgA. Among most of the subjects who received two or three vaccine doses, the detected levels of Spike-specific s-IgA responses were either below the cut-off of positivity or they were very low (24 out of 29 double-vaccinated and 18 of 25 in triple-vaccinated below the cut-off), whereas previous infection elicited significantly higher anti-triS s-IgA responses in comparison to vaccination (15 out of 21 above cut-off) (**Figure 2C**). Similar to anti-triS IgA responses, there also was no significant difference of anti-triS s-IgA between vaccinated subjects which received two or three vaccine doses, indicating that a 3^rd^ vaccine dose does not boost local s-IgA responses.

We then analyzed the influence of vaccination and infection on mucosal and systemic IgG responses. We detected significantly higher anti-triS IgG responses in NLF of vaccinated participants compared to the convalescent group, while among vaccinated individuals higher anti-triS IgG titers were found in subjects who received 3 vaccine doses (**Figure 2D**). A similar pattern was observed in serum anti-triS IgG responses, confirming the highly similar profiles of SARS-CoV-2 IgG responses in nasal mucosa and serum (**Figure 2E**).

### Influence of hybrid immunity on mucosal antibody responses

To evaluate the impact of hybrid immunity on mucosal antibody responses, we compared antibody titers between convalescent participants and vaccinated participants which were subsequently infected. We detected moderately higher anti-triS IgA responses in NLF of individuals with hybrid immunity in comparison to convalescent subjects, however these were significantly higher only in subjects that have received three vaccine doses (**Figure 3A**). Serum anti-triS IgA responses were significantly higher in subjects with hybrid immunity after 1/2 or 3 doses in comparison to convalescent subjects (**Figure 3B**). To assess the influence of hybrid immunity on the locally-generated immune responses, we determined the levels of anti-triS s-IgA in these groups. Moderately elevated levels of anti-triS s-IgA were detected in individuals with hybrid immunity compared to convalescent subjects, however these were only significant in subjects that received 1/2 vaccine doses **(Figure 3C**). These results suggest that vaccination before infection has only a moderate impact on locally-generated immune responses.

**Figure 3.**
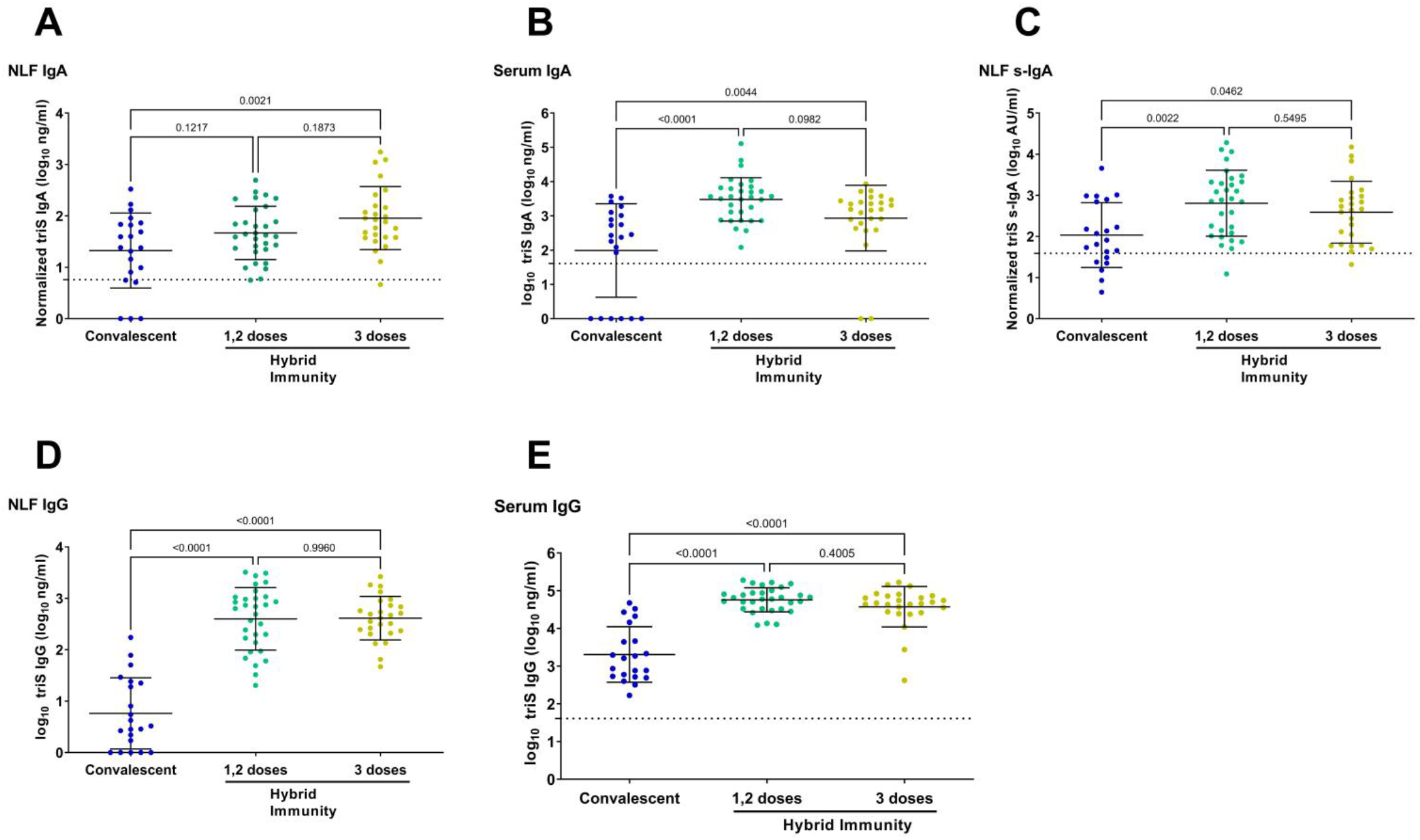
Mucosal and serum antibody responses in convalescent compared to subjects with hybrid immunity. A, B. Anti-triS IgA (log_10_ ng/ml) measured in NLF (A) and serum (B) of convalescent or subjects with hybrid immunity. (C) Anti-triS s-IgA (log_10_ AU/ml) antibody titers in NLF of convalescent or subjects with hybrid immunity. D,E. Anti-triS IgG measured in NLF (D) and serum (E) of convalescent or subjects with hybrid immunity. One-way ANOVA with Tukey test was used to determine differences of means, p values are shown above brackets. The mean + 3SD of 11 negative samples was used to set a cut-off for anti-triS IgA, s-IgA and IgG in NLF. The mean + 3SD of 56 pre-pandemic serum samples was used to set a cut-off for anti-triS IgA in serum samples. The mean + 3SD of 48 pre-pandemic serum samples was used to set a cut-off for anti-triS IgG in serum samples.

We then evaluated the impact of hybrid immunity on local and systemic IgG responses. Anti-triS IgG responses in NLF were significantly higher in vaccine breakthroughs in comparison to convalescent participants (**Figure 3D**). Similar to NLF, anti-triS IgG serum responses were also significantly elevated in breakthroughs in comparison to convalescent participants (**Figure 3E**). These results suggest that while vaccination boosts local IgG responses, it has rather limited impact on local IgA responses.

We also examined if anti-NP s-IgA responses are induced by infection in NLF and compared them to anti-triS antibody titers. Remarkably, there was only weak correlation between anti-triS and anti-NP s-IgA mucosal responses in individuals with previous infection (Pearson r=0.19; p=0.1) (**Figure S3A**). In the majority of subjects with previous infection anti-NP s-IgA responses in NLF were below the cut-off of positivity (17 out of 21 in convalescent, 24 out of 31 in subjects with hybrid immunity after 1/2 doses and 17 of 26 in subjects with hybrid immunity after 3 doses and) and there was no significant difference between convalescent subjects and vaccine breakthroughs (**Figure S3B**), suggesting that anti-NP s-IgA responses are either produced at very low levels or wane quickly.

### Neutralization capacity of mucosal antibodies

Last, we asked how efficient mucosal antibodies neutralize the ancestral SARS-CoV-2 strain and the Omicron BA.5 variant. Therefore, we performed a focus reduction neutralization test (FRNT) using NLF from participants that were only vaccinated with three vaccine doses and subjects with hybrid immunity acquired either by infection with the Delta or Omicron BA.1 variant after vaccination. Individuals with hybrid immunity had significantly higher neutralizing antibody titers against the ancestral SARS-CoV-2 strain in NLF compared to vaccinated individuals independently of the infecting variant (**Figure 4A left panel**). Next, we wanted to assess whether individuals with Delta and Omicron BA.1 derived hybrid immunity were better protected against infection with Omicron BA.5 compared to vaccinated subjects. We observed significantly higher neutralization capacity against Omicron BA.5 in subjects that were previously infected with Omicron BA.1 (**Figure 4B, left panel**). Since neutralization is measured isotype independent in our assay, we wanted to determine the contribution of mucosal s-IgA. Therefore, we correlated neutralizing antibody titers against the ancestral strain and Omicron BA.5 with s-IgA binding antibodies directed against triS. Anti-triS s-IgA binding antibodies in NLF showed a moderate to strong correlation with neutralizing activity against both SARS-CoV-2 strains (**Figure 4A and B, right panel**). These findings indicate that s-IgA is a pivotal contributor to neutralization in the mucosa.

**Figure 4.**
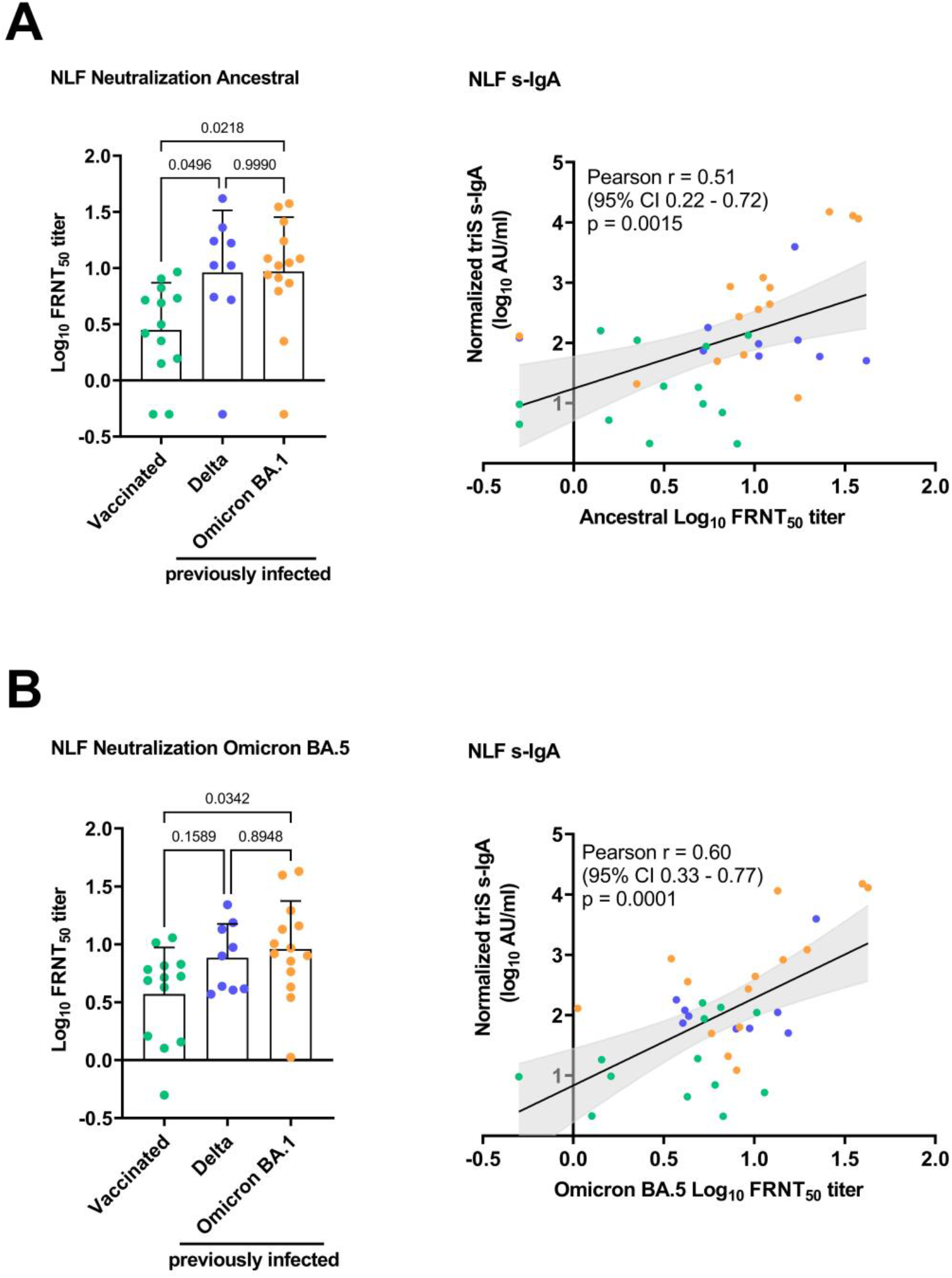
Neutralizing antibody responses in the nasal mucosae of vaccinated and subjects with hybrid immunity. (A) Each dot represents the neutralizing titer (FRNT_50_) of an individual NLF sample against SARS-CoV-2 ancestral (A) and Omicron BA.5 variant (B). One-way ANOVA with Tukey test was used to determine differences of means, p values are shown above brackets. Anti-triS s-IgA and IgG titers in NLF from the same subjects were correlated with FRNT_50_ neutralizing titers against SARS-CoV-2 ancestral (A) and Omicron BA.5 variant (B). Pearson correlation coefficient and p values are shown.

## Discussion

In this study, we investigated mucosal antibody responses following COVID-19 vaccination, infection with SARS-CoV-2 or infection in vaccinated individuals, i.e. hybrid immunity. We demonstrated that s-IgA antibodies, which are present only on mucosal surfaces and not in systemic circulation, are the main contributor to neutralization in the mucosa. Additionally, we have shown that s-IgA responses are significantly elevated in subjects with previous infection, whereas vaccination-induced s-IgA responses are rarely detected and only at a very low level. Moreover, we found significantly higher neutralization titers against the ancestral SARS-CoV-2 strain in individuals with hybrid immunity compared to vaccinated subjects, providing evidence that previous infection elicits more potent functional mucosal antibodies. Interestingly, infection with Omicron BA.1 or Delta led to a similar neutralization capacity against Omicron BA.5 despite the smaller antigenic distance between BA.1 and BA.5, suggesting that the infecting variant is less important for protection. Contrary, in a study by Malato et al. a higher protection against BA.5 infection was found in individuals previously infected with BA.1/2 compared to Delta. However, in this study the difference in protection from Omicron BA.5 infection by previous infection with BA.1/2 or Delta was small and potential increased waning of immunity in Delta infected participants due to the longer time span was not accounted for^30^.

Our results show that mucosal and systemic IgG responses are highly similar, whereas mucosal IgA responses are compartmentalized from systemic responses. This is consistent with previous studies that show the discordance of local and systemic SARS-CoV-2 antibody responses^15,16,31^. In convalescent subjects, neutralization activity poorly correlated between nasopharyngeal and blood samples ^16,31^. Interestingly, potent antibody responses in nasal fluids were found in some seronegative participants^15,16^, indicating that in some cases SARS-CoV-2 infections lead only to a local but no systemic response.

We demonstrated that currently available intramuscularly-administered vaccines have a limited impact on SARS-CoV-2 specific mucosal responses. These results go in line with another study showing that vaccination efficiently boosts nasal IgG responses whereas nasal IgA responses are only transiently increased, and rapidly decline after vaccination^19^. In another study vaccination did not generate detectable neutralizing mucosal antibodies and only breakthrough infections in vaccinated subjects resulted in measurable neutralization^28^. To date, the presence of SARS-CoV-2 s-IgA responses were detected in saliva and breastmilk of some individuals upon vaccination^32^. However, the levels of s-IgA antibodies in most subjects, who received mRNA vaccines was very low or below the detection limit^26^. According to one study, anti-triS s-IgA responses were detected in saliva in 30% of subjects after two doses of mRNA vaccines, and the detected levels were significantly lower compared to convalescent patients^25^. It remains unknown how mucosal s-IgA responses can be induced upon intramuscular vaccination. SARS-CoV-2 spike protein was detected in plasma after mRNA vaccination, and the clearance of this antigen correlated with the production of IgA antibodies, indicating that this antigen could reach the MALT to further induce mucosal antibody responses^33^. Alternatively, after vaccination antigen diffuses to the regional draining lymph nodes, where it is taken up by local antigen-presenting cells, that can further migrate to the MALT and activate B cells that generate s-IgA antibodies^34^.

Here we show that s-IgA is an important contributor to neutralization in the NLF. Similarly, it has been demonstrated that IgA antibodies in nasal secretions are most strongly correlated with SARS-CoV-2 neutralization^35^. Another study has shown that depletion of IgA from nasal wash samples lead to a reduction of the neutralization capacity^31^. One study demonstrated that higher levels of mucosal IgA but not IgG antibodies correlated with lower levels of viral replication and lower risk of infection with Omicron^36^. Furthermore, increased disease severity and mortality was identified among patients with IgA deficiency^37^. However, further studies which would evaluate the role of pre-existing immunity on SARS-CoV-2 shedding and infection rates are currently missing. A recent study evaluated the effect of different vaccine administration routes on viral transmission in hamsters. Intranasally-administered adenovirus-vector vaccine or infection lead to significantly lower cumulative shedding or airborne transmission in comparison to intramuscular vaccine administration^38^. Indeed, mucosally-administered vaccines are proposed as a strategy to reduce onward transmission. In our study, we detected the most potent local and systemic antibody responses in subjects with hybrid immunity. Heterologous vaccination strategies that mimic hybrid immunity by giving a systemic prime through intramuscular vaccination followed by a mucosal boost, by intranasal vaccination, were shown to induce robust T and B cell immunity in the respiratory mucosa^39,40^. It is currently unknown how long mucosal antibodies titres induced by mucosal vaccination remain sufficiently elevated to protect from SARS-CoV-2 infection. A recent study demonstrated that protection mediated by mucosal IgA antibodies lasted at least for 8 months following SARS-CoV-2 infection^10^.

This study has some limitations. All participants were sampled only at a single time point, therefore we compared antibody responses between different individuals rather than following the same individuals to compare changes of antibody responses over time or after subsequent immune reactions. Moreover, the sampling time points were not identical for all the participants within each group. However, all groups have similar age and sex distribution validating our conclusions. Most of the study participants were young adults, which might present more potent immune responses in comparison to older subjects, therefore these results cannot be extrapolated across age groups. Furthermore, all of the participants received mRNA vaccines, while mucosal antibody responses to other types of COVID-19 vaccines, mainly used in the low- and middle-income countries, have not yet been investigated.

While serum neutralization titers are highly predictive of immune protection from COVID-19 disease, correlates of protection from infection and transmission are not well defined. In this study we have shown that prior infection leads to more robust mucosal binding and neutralizing antibody responses, with s-IgA seemingly to play a crucial role. Therefore, the development of vaccines that elicit strong and lasting mucosal antibody responses would be vital to curtail infectious shedding and further transmission.

## Acknowledgements

We thank Pauline Vetter, Morgann Duverger, Rachel Goldstein and Christiane Eberhardt for their help with clinical sample collection. We thank Florence Pojer, Kelvin Lau, and David Hacker from the EPFL Protein Production Facility for providing us with the purified SARS-CoV-2 spike protein.We thank the staff of the laboratory of virology at the HUG for support. We are grateful for the patients who were willing to donate their samples and agree to participate in our research.

## Funding

The work was funded by the COVID-19 National Research Program (grant number 198412) of the Swiss National Science Foundation.

## Author contributions

O.P., I.E. and B.M. conceptualized the study. O.P., K.H.F., Ma.B. and Me.B. conducted the recruitment of participants. O.P., Ma.B., K.H.F. and Me.B. collected the clinical samples. O.P., K.A, Me.B. P.S., S.C.B, I.A.V performed the laboratory experiments. O.P., I.E. and B.M. analyzed and interpreted the data. I.E. and B.M. supervised the work. O.P., I.E. and B.M. wrote the manuscript. All authors contributed to the final draft.

## Supplementary material

**Table S1.**
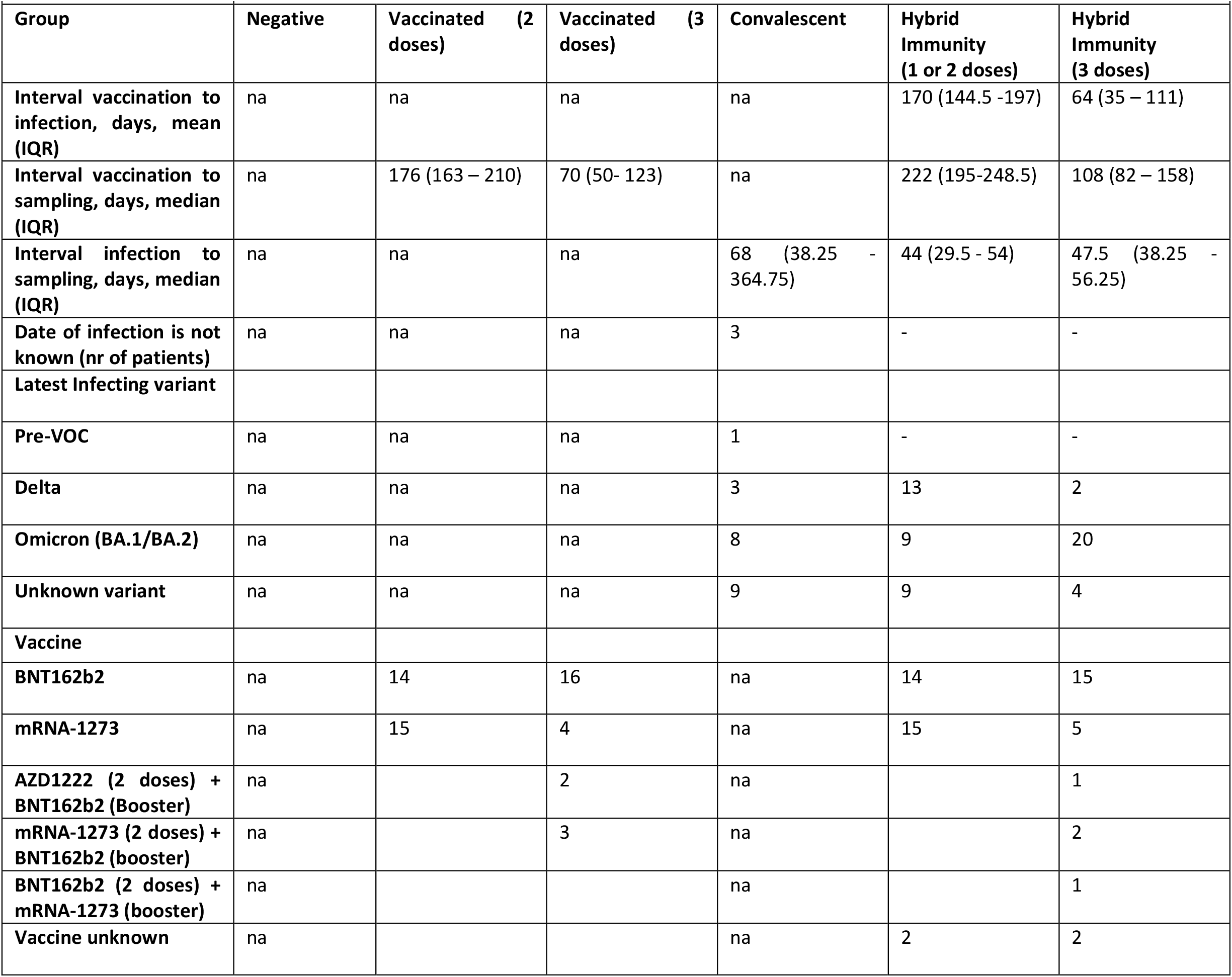
Participants characteristics (extended)

**Figure S1.**
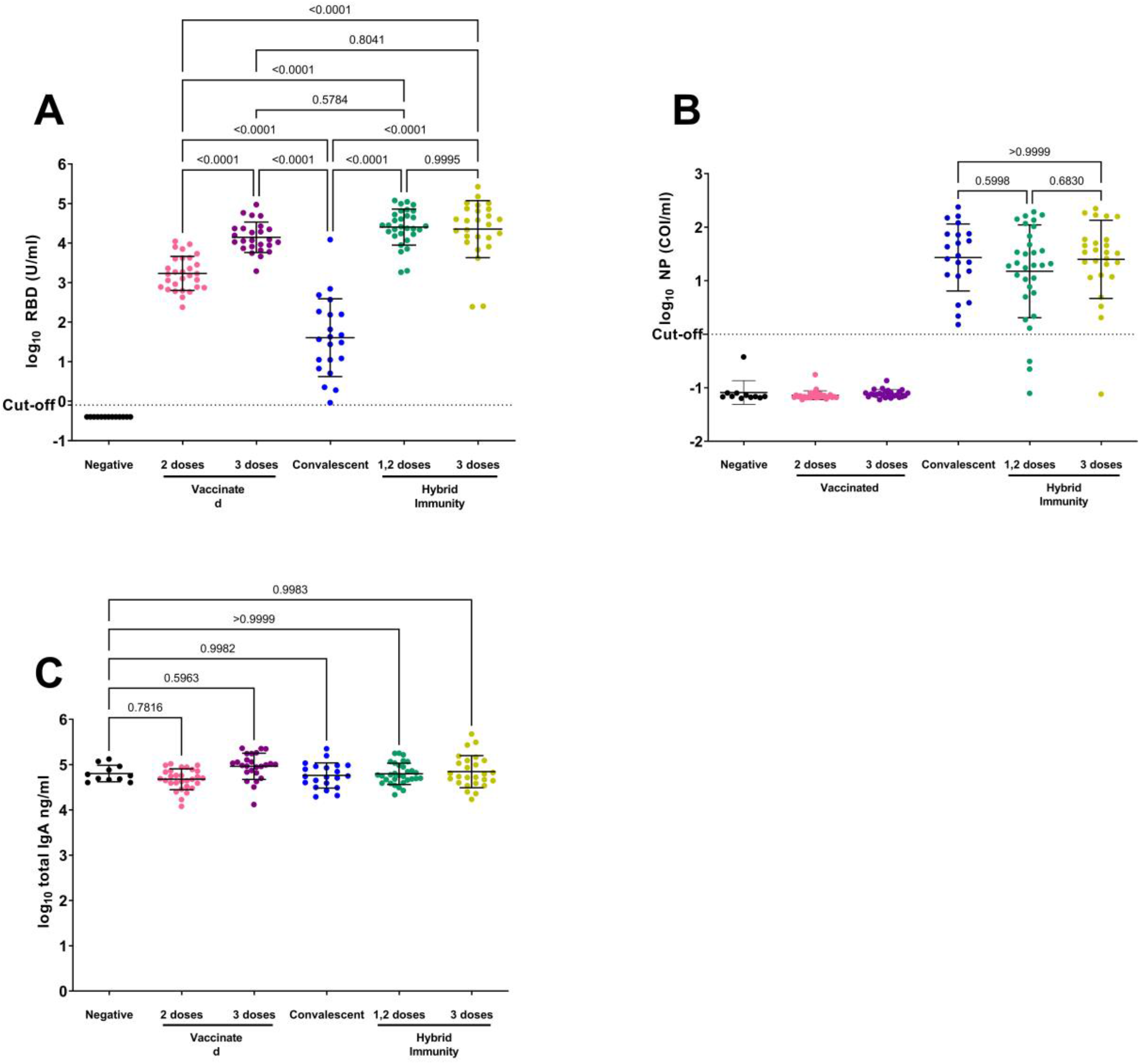
Validation of serum and NLF samples. The levels of SARS-CoV-2 anti-RBD (A) and NP (B) were measured in serum using Roche Elecsys anti-SARS-CoV-2 S (A) and SARS-CoV-2 (B) total Ig assay. (C) Levels of total IgA in the NLF samples. One-way ANOVA with Tukey test was used to determine differences of means, p values are shown above brackets.

**Figure S2.**
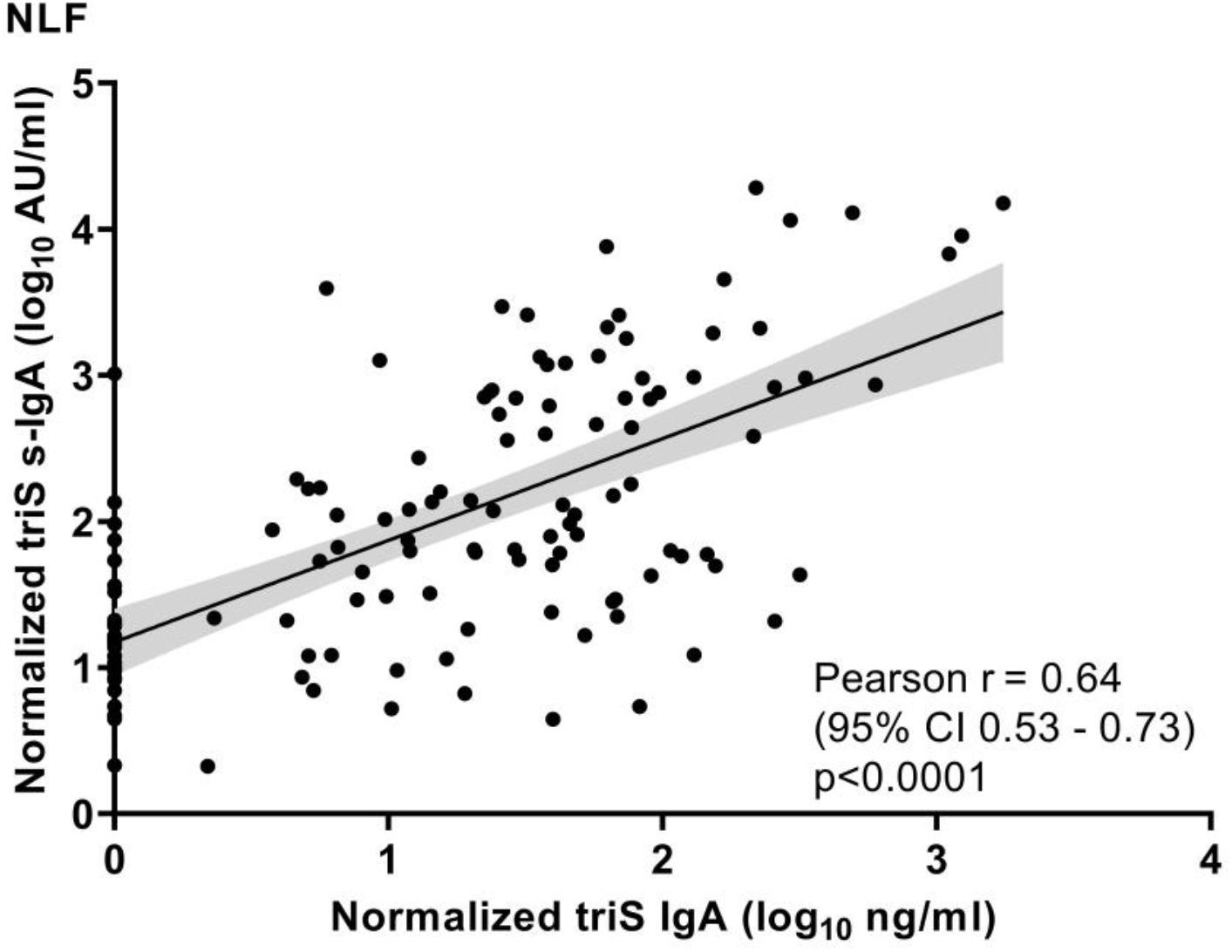
SARS-CoV-2 anti-Spike s-IgA and IgA responses in nasal mucosa. Correlation between anti-triS IgA and s-IgA antibody titers in NLF collected from convalescent, vaccinated and subjects with hybrid immunity. Pearson correlation coefficient and p values are shown.

**Figure S3.**
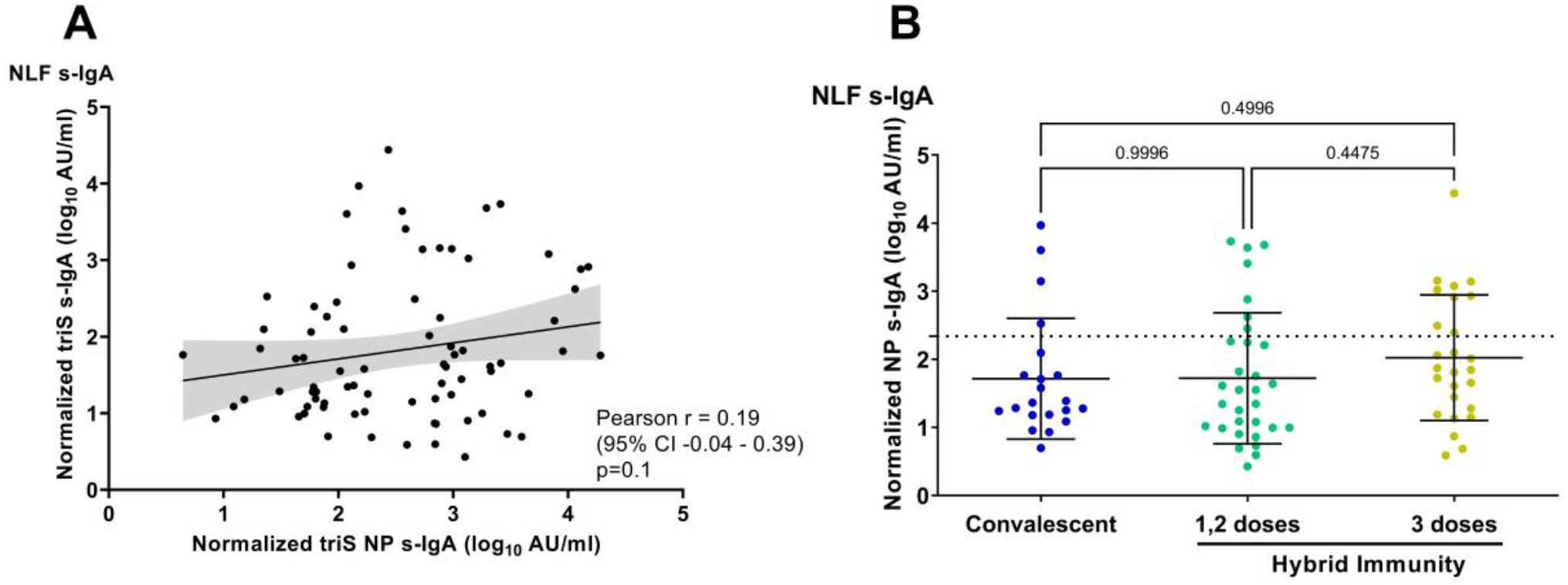
SARS-CoV-2 anti-nucleoprotein responses in individuals with previous infection. (A) Correlation between anti-triS and anti-NP s-IgA titers in NLF samples of convalescent or individuals with hybrid immunity. Pearson correlation coefficient and p values are shown. (B) Levels of anti-NP s-IgA titers of convalescent or individuals with hybrid immunity. One-way ANOVA with Tukey test was used to determine differences of means, p values are shown above brackets.

## Notes

### Competing Interest Statement

The authors have declared no competing interest.

### Author Declarations

The study was approved by the Cantonal Ethics Committee at the University Hospital of Geneva (CCER no. 2020-02323).

